# Estimating the Global Market Size for Disease-Modifying Therapies in Epilepsy Caused by Haploinsufficiency

**DOI:** 10.1101/2025.01.20.25320871

**Authors:** Eduardo Pérez-Palma, Tobias Brünger, Samden Lhatoo, Guo-Qiang Zhang, Fabio A Nascimento, Dennis Lal

## Abstract

**Importance:** Haploinsufficiency, a condition where a single functional copy of a gene is insufficient to maintain normal physiological function due to a mutation, represents the most prevalent disease mechanism underlying genetic epilepsies, particularly developmental and epileptic encephalopathies (DEEs). Current therapies for these patients primarily address symptoms and fail to alter the disease trajectory. Given the prospect of targeted gene therapies, accurately quantifying the burden of these disorders is paramount for optimizing resource allocation in patient care and facilitating the development of novel disease-modifying therapies.

**Objective:** To estimate the global and U.S. incidence and prevalence of epilepsy caused by haploinsufficiency.

**Design, Setting, and Participants:** First, we identified epilepsy genes, focusing on autosomal dominant genes with a high likelihood of haploinsufficiency. Second, we utilized a statistical framework to estimate the birth incidence of patients with protein-truncating variants in these genes, incorporating gene-specific mutation rates and birth rate data. Finally, we calculated prevalence by multiplying annual incidence by an effective disease duration, considering factors like sudden unexpected death in epilepsy.

**Main Outcomes and Measures:** Birth incidence and prevalence of epilepsy attributable to *de novo* protein-truncating variants in haploinsufficient genes.

**Results:** An estimated 228.81 (90% CI: 206.21–241.93) cases of haploinsufficiency-related epilepsy occur per 100,000 live births globally. This translates to approximately 313,595 new cases annually worldwide and 8,736 in the United States. The estimated global prevalence is 14,343,840 individuals.

**Conclusions and Relevance:** This study provides the first data-driven estimates of the of haploinsufficiency-related epilepsy burden, highlighting a substantial population potentially benefiting from gene-targeting therapies. These findings have significant implications for research prioritization, resource allocation, and the development of precision medicine approaches for treating these severe neurological disorders.

## Introduction

Developmental and epileptic encephalopathies (DEEs)^1^ represent rare and severe epilepsy syndromes marked by numerous comorbidities, both neurological and non-neurological^2,3^. Most DEEs are sporadic, monogenic and caused by spontaneous single ‘*de novo’* variants, only present in the affected patient^4^. In DEEs, no current drugs can significantly improve the development of affected patients and, as a result, they often suffer from lifelong epilepsy and disabilities. Thus, the prospect of targeted therapies—such as gene replacement and upregulation strategies—has garnered significant interest^5^. Most genetic epilepsies relates to haploinsufficiency, where the non-mutated gene copy produces insufficient functional protein to compensate for the mutated one. For most haploinsufficient genes, disease is predominantly caused by protein-truncating variants, which are straightforward to interpret clinically. This clarity facilitates precision medicine approaches, including confident genetic testing interpretation in the clinical care and strategies to design precision therapies to enhance protein production from the functional gene copy or introduce a non-mutated protein through gene replacement therapy.^6^

Haploinsufficiency is not universally implicated with all epilepsy-associated genes, and epidemiological data specific to haploinsufficiency-related epilepsies remains unavailable^4,7^. This lack of data hampers efforts to quantify the disease burden, impeding the allocation of healthcare resources, the design of targeted treatment programs, and the evaluation of novel gene-based therapies. These gaps also challenge decision-making within pharmaceutical and investment sectors, where reliable epidemiological information is essential for market sizing, demand forecasting, and research prioritization^5,6^. Building on well-established prior work on genetic-informed incidence estimates by our team ^8^and others^9^, we provide the first comprehensive estimates of the incidence and prevalence of haploinsufficiency-related epilepsy. Using mutation rate modeling, loss-of-function intolerance metrics, and ethnicity-specific birth rate data, we estimate for the first time the global and U.S. burden of these genetic epilepsies.

## Methods

### Gene Selection and Characterization

We selected epilepsy-associated genes using an expert-curated repository (Genes4Epilepsy)^4^, focusing on autosomal dominant genes with a loss-of-function (LoF) observed-to-expected ratio ≤0.35^10^ to capture likely haploinsufficiency (see eMethods for details).

### Mutation Rate and Birth Incidence Calculations

We estimated birth incidence for haploinsufficiency-related epilepsy using a framework adapted from Samocha et al.^9^ and López-Rivera et al.^8^, restricting our analysis to protein-truncating variants (PTVs). Because most DEEs arise *de novo*, we calculated per-gene rates of deleterious PTVs. For eight genes lacking published mutation rates, we imputed values based on coding gene length, which correlates with PTV frequency (see eMethods).

### Global regions and US ethnicity-Specific Analysis

We estimated incidences using births rates from world bank data.^11^ To explore differences in U.S. populations, we stratified incidence estimates by ethnic birth rates NCHS National Health Statistics Reports^12^. Smaller groups (e.g., American Indian/Alaska Native) were also included (see eMethods).

### Prevalence Estimation

We derived prevalence by multiplying annual incidence by effective disease duration, incorporating data on life-years lost for individuals with early-onset epilepsy and SUDEP-related deaths (see eMethods).

### Statistical and Computational Framework

All analyses were conducted in R (v4.3), employing custom scripts for incidence calculations, regression models, and imputation checks (see eMethods).

### Results

Our primary objective was to determine the proportion of epilepsy cases caused by *de novo* pathogenic variants leading to haploinsufficiency. First, we identified “well-established” epilepsy genes using the *Genes4Epilepsy* resource, prioritizing autosomal dominant genes that are strongly constrained for protein-truncating variants (see eMethods). This approach yielded 289 genes, 90.1% of which are associated with DEEs.

We next modeled the birth incidence of *de novo* protein-truncating variants (PTVs) in these genes, focusing on straightforwardly interpretable truncating events (see Methods). Across the 289 epilepsy-associated genes, we calculated a total aggregated annual birth incidence of 228.81 (90% CI: 206.21–241.93) per 100,000 live births for haploinsufficiency-associated epilepsy. This translates to an estimated annual global incidence of 313,595 new cases (global birth rate: 17/1,000 live births) and 8,736 new cases in the United States (U.S. birth rate: 11.4/1,000 live births) attributable to truncating variants. Notably, 72 genes (25%), accounted for a >50% proportion of the estimated incidence.

Subsequent analyses examined incidence by ethnicity within the United States, highlighting the impact of varying population sizes and birth rates (see Methods). Among these groups, the US White population (57.8%) has the highest number of annual new cases (4,846), reflecting its large population base. However, ethnic groups with higher natality rates, such as Hispanic or Latina (18.7%), have a proportionally higher incidence per 100,000 births, leading to 1,952 new cases annually despite representing a smaller population share. Similarly, Black or African American (12.1%) and Asian or Pacific Islander (5.9%) populations show consistent patterns of new cases relative to their population sizes and birth rates, with 784 and 590 annual new cases, respectively. The American Indian or Alaska Native (0.7%) group exhibits the lowest total cases (47 annually), reflecting both a smaller population and a lower natality rate.

Finally, to estimate prevalence, we multiplied the annual incidence by an effective disease duration that factors in DEE-related mortality and SUDEP (see Methods). Given that over 90% of associated genes (see previous section) are linked to DEEs, the genetic disorder is expected to persist lifelong. To estimate life expectancy in this population, we applied a previously published value for life-years lost in individuals with epilepsy before age 15 (average 63.45 years)^13^, adjusted for an average SUDEP rate of 6.1 per 1,000 person-years in individuals with DEEs^3^. This adjustment yielded an effective disease duration, equivalent to life expectancy, of 45.74 years. Using this estimate, we calculated a global prevalence of approximately 14,343,840 individuals and a U.S. prevalence of 399,595 individuals with haploinsufficiency-associated genetic epilepsies.

## Discussion

Our findings offer a critical data-driven baseline reference in quantifying haploinsufficiency-related epilepsies, highlighting a substantial yet previously undefined population of patients who may benefit from gene replacement or upregulation therapies. By focusing on protein-truncating variants—overwhelmingly implicated in haploinsufficiency—we address a mechanistic, rather than purely phenotypic, approach to incidence modeling. This strategy mirrors successful applications in precision oncology, where genetically defined subgroups enabled targeted treatments and data-driven investment^14,15^.

Drawing on curated genetic resources^4^ and well-established mutational frameworks^8,9^, our estimates illustrate both the clinical and commercial significance of gene-based interventions. While the application of mutation rate models and intolerance metrics provides a sound basis for global and U.S. incidence and prevalence projections, several caveats warrant caution. First, restricting analyses to *de novo* truncating variants likely underestimates the total burden of haploinsufficiency, as missense, X-linked, inherited, or copy number variants were excluded. Second, epilepsy diagnosis, SUDEP rates and life-years lost are influenced by penetrance and diagnostic variability which can also lead to under ascertainment of affected individuals.

Despite these limitations, our work highlights the remarkable lifelong burden associated with haploinsufficiency-related DEEs. Such data may galvanize therapeutic development by clarifying potential patient populations and encouraging biomarker-driven clinical trials. Our prevalence estimates also highlight the genetic diagnosis gap in adult patients due to historical cost barriers, the scarcity of adult genetic centers, and financial or logistical challenges. Further, improved care has extended the lifespan of DEE patients, amplifying the lifelong burden on patients, caregivers, and healthcare systems. Here, optimized clinician education will enhance diagnosis, management, and the implementation of emerging genetic therapies.

Understanding the substantial number of individuals affected provides critical context for regulatory planning and payor considerations, potentially expediting access to novel therapies. Future efforts should focus on validating these estimates through population-based registries and expanding them to include non-truncating and inherited variants.

## Supporting information

eMethods

## Data Availability

All data produced in the present work are contained in the manuscript

## References

1. Scheffer IE, Berkovic S, Capovilla G, et al. ILAE classification of the epilepsies: Position paper of the ILAE Commission for Classification and Terminology. Epilepsia. 2017;58(4):512–521. doi:10.1111/epi.13709

2. Scheffer IE, Zuberi S, Mefford HC, Guerrini R, McTague A. Developmental and epileptic encephalopathies. Nat Rev Dis Primer. 2024;10(1):1–19. doi:10.1038/s41572-024-00546-6

3. Donnan AM, Schneider AL, Russ-Hall S, Churilov L, Scheffer IE. Rates of Status Epilepticus and Sudden Unexplained Death in Epilepsy in People With Genetic Developmental and Epileptic Encephalopathies. Neurology. 2023;100(16):e1712–e1722. doi:10.1212/WNL.0000000000207080

4. Oliver KL, Scheffer IE, Bennett MF, Grinton BE, Bahlo M, Berkovic SF. Genes4Epilepsy: An epilepsy gene resource. Epilepsia. 2023;64(5):1368–1375. doi:10.1111/epi.17547

5. Klein P, Kaminski RM, Koepp M, Löscher W. New epilepsy therapies in development. Nat Rev Drug Discov. 2024;23(9):682–708. doi:10.1038/s41573-024-00981-w

6. Evans EF, Shyr ZA, Traynor BJ, Zheng W. Therapeutic development approaches to treat haploinsufficiency diseases: restoring protein levels. Drug Discov Today. 2024;29(12):104201. doi:10.1016/j.drudis.2024.104201

7. Carvill GL, Matheny T, Hesselberth J, Demarest S. Haploinsufficiency, Dominant Negative, and Gain-of-Function Mechanisms in Epilepsy: Matching Therapeutic Approach to the Pathophysiology. Neurotherapeutics. 2021;18(3):1500–1514. doi:10.1007/s13311-021-01137-z

8. López-Rivera JA, Pérez-Palma E, Symonds J, et al. A catalogue of new incidence estimates of monogenic neurodevelopmental disorders caused by de novo variants. Brain. 2020;143(4):1099–1105. doi:10.1093/brain/awaa051

9. Samocha KE, Robinson EB, Sanders SJ, et al. A framework for the interpretation of de novo mutation in human disease. Nat Genet. 2014;46(9):944–950. doi:10.1038/ng.3050

10. Karczewski KJ, Francioli LC, Tiao G, et al. The mutational constraint spectrum quantified from variation in 141,456 humans. Nature. 2020;581(7809):434–443. doi:10.1038/s41586-020-2308-7

11. World Bank Open Data. World Bank Open Data. Accessed December 22, 2024. https://data.worldbank.org

12. Martin JA, Hamilton BE, K OMJ, Driscoll AK. Births: Final Data for 2019. Accessed December 22, 2024. https://stacks.cdc.gov/view/cdc/100472

13. Dreier JW, Laursen TM, Tomson T, Plana-Ripoll O, Christensen J. Cause-specific mortality and life years lost in people with epilepsy: a Danish cohort study. Brain J Neurol. 2023;146(1):124–134. doi:10.1093/brain/awac042

14. Marquart J, Chen EY, Prasad V. Estimation of the Percentage of US Patients With Cancer Who Benefit From Genome-Driven Oncology. JAMA Oncol. 2018;4(8):1093–1098. doi:10.1001/jamaoncol.2018.1660

15. Haslam A, Prasad V. Estimation of the Percentage of US Patients With Cancer Who Are Eligible for and Respond to Checkpoint Inhibitor Immunotherapy Drugs. JAMA Netw Open. 2019;2(5):e192535. doi:10.1001/jamanetworkopen.2019.2535

