## Supplementary material for "Estimating the Global Market Size for Disease-Modifying Therapies in Epilepsy Caused by Haploinsufficiency": eMethods

### Supplemental Online Content (eMethods)

Eduardo Pérez-Palma PhD[1], Tobias Brünger PhD[2], Samden Lhatoo MD[2], Guo-Qiang Zhang PhD[2], Fabio A Nascimento MD[3], Dennis Lal PhD[2,4,5]

[1] Universidad del Desarrollo, Centro de Genética y Genómica, Facultad de Medicina Clínica Alemana, Santiago, Chile.

[2] Department of Neurology, McGovern Medical School, The University of Texas Health Science Center at Houston, 1133 John Freeman Blvd, Houston, TX 77030, United States.

[3] Department of Neurology, Washington University School of Medicine, St. Louis, Missouri, U.S.A.

[4] Program in Medical and Population Genetics, Broad Institute of Massachusetts Institute of Technology (M.I.T.) and Harvard, 415 Main St, Cambridge, MA 02142, United States.

[5] Stanley Center for Psychiatric Research, Broad Institute of Harvard and M.I.T, 415 Main St., Cambridge, MA 02142, United States.

**eMethods.**

This supplemental material has been provided by the authors to give readers additional information about their work.

**Rationale and strategic Approach.**

Here, we provide an overview and rational of the strategic approach: Despite decades of research, clinical interventions for developmental and epileptic encephalopathies (DEEs) remain largely symptomatic and descriptive, highlighting the urgent need for therapies that directly address the underlying genetic mechanisms by which the disease arise[1]. Our study aims to quantify the global and U.S. burden of haploinsufficiency-related epilepsies—conditions primarily driven by loss-of-function variants—thus laying the groundwork for precision treatments such as gene replacement or upregulation. To achieve this, we leveraged the Genes4Epilepsy resource[2], which provides a rigorously curated set of epilepsy genes and evidence-based classifications (see eMethods below). We narrowed our focus to autosomal dominant genes with a loss-of-function observed-to-expected ratio (LoF o/e) ≤0.35, a threshold established by population-genomic data from Karczewski *et al*[3], to capture only those genes most likely to exhibit haploinsufficiency. We then harnessed a statistical framework, initially proposed by Samocha *et al*.[4] and further extended by López-Rivera et al.[5], to estimate the incidence of deleterious protein-truncating variants (PTVs)—a conservative but clinically interpretable class of mutations that frequently leads pathogenic haploinsufficiency. Our framework is based on simple idea: The difference between the number of observed vs expected PTVs in the general population should be the estimated number of pathogenic PTVs in the general population (López-Rivera, 2014). These enabled us to calculate the aggregated total incidence of all monogenic haploinsufficient epilepsies.

In parallel, we integrated global[6] and ethnicity-specific birth rate data to explore potential disparities across different U.S. populations[7], providing a more nuanced picture of disease incidence. We also computed prevalence by factoring in disease duration estimates[8], including considerations for SUDEP and non-specific mortality often associated with DEEs[9]. Finally, we describe our analytical pipeline—which includes custom R scripts for incidence modeling, linear regressions, and imputation procedures—in detail, ensuring transparency and reproducibility for future investigators. This comprehensive approach offers the first robust epidemiological assessment of haploinsufficiency-related epilepsy and informs the development of targeted gene-based therapies.

**Detailed Methods.**

**Gene Selection and Characterization.**

We utilized the *Genes4Epilepsy* resource[2], an expert-curated compilation updated biannually, to identify genes associated with epilepsy. The curation process incorporates robust evidence-based criteria, including pathogenicity confirmation and relevance to epilepsy phenotypes, ensuring high reliability. For this study, we accessed the resource on December 2024 that included 1016 genes. From an initial list, we prioritized genes classified as autosomal dominant and filtered in those with a loss-of-function observed-to-expected (LoF o/e) ratio ≤0.35 to only keep genes likely to exhibit haploinsufficiency[3].

**Mutation Rate and Birth Incidence Calculations.**

We used a statistical framework originally described by Samocha et al.[4] and extended by López-Rivera et al.[5] to estimate birth incidence for haploinsufficiency-related epilepsy. Because most developmental and epileptic encephalopathies (DEEs) arise sporadically from *de novo* variants—particularly among the 289 autosomal-dominant genes identified—we calculated the frequency of these variants per gene and birth based on well-established mutation rates generated from large genomic population datasets[4]. Focusing on protein-truncating variants (PTVs) provides a conservative but straightforward estimate of disease burden[5]. This method combines gene-specific PTV mutation rates (nonsense, frameshift, splice-site) with intolerance metrics to approximate the likelihood of deleterious de novo mutations. Local sequence context and gene length, known to influence mutability, were factored into these calculations. For eight genes (2.77%, 8/289) lacking published mutation rate data (*AFDN*, *ADGRL1*, *KMT5B*, *MARCHF6*, *PHACTR1*, *CNOT9*, *SCAMP5*, *H3-3B*), we used linear regression based on coding gene length to impute missing values. Gene length has proven to be a robust proxy for estimating the number of expected PTVs, thereby ensuring a consistent and reliable approach to incidence modeling. Empirical correlation between base pairs and PTV mutation rates resulted in an $R^2$ value of 0.978.

**Ethnicity-Specific Analysis.**

To examine disparities across U.S. ethnic groups, we incorporated birth rate stratifications by ethnicity from national statistics[7]. Incidence was proportionally adjusted based on ethnic-specific natality data. For smaller populations (e.g., American Indian/Alaska Native).

**Prevalence Estimation.**

Prevalence was calculated by multiplying annual incidence of new cases by effective disease duration. Disease duration value for Haploinsufficient Monogenic Epilepsies was derived from life-years lost data of people developing epilepsy before 15 years old[8] and incorporated developmental epileptic encephalopathy related unspecific deaths and SUDEP[9].

**Statistical and Computational Framework.**

Analyses were conducted using R (v4.3), employing custom scripts for incidence calculations and regression modeling. Key statistical tests included Pearson correlation for imputation validation.

**Considerations related to developmental disorders beyond epilepsy.**

According to the Centers for Disease Control and Prevention (CDC), during 2021 and 2022, about 2.9 million U.S. adults aged 18 and older reported having active epilepsy, which is approximately 1% of all U.S. adults[10]. If one considers that DEEs are a subset of all cases of epilepsy, our prevalence estimations of 399,595 DEE cases represents approximately 11.75% of the total population with epilepsy in the U.S. This might be considered an overestimation, however is not. Some genes traditionally associated with Developmental and Epileptic Encephalopathies (DEE) can also cause neurodevelopmental disorders without epilepsy[11], highlighting their pleiotropic nature. For example, mutations in genes like *SCN2A*[12], *SCN8A*[13], and *TSC*[14] have been linked to conditions such as intellectual disability, developmental delays, and autism spectrum disorders, even in the absence of seizures. Thus, our gene-based calculations highlight total disease prevalence due to haploinsufficiency within these genes, regardless if seizures are present. Further, our estimations accounts for all individual who might benefit from gene-therapy. This broadens the understanding of the impact these genes have on neurodevelopmental phenotypes, emphasizing the need for a gene-oriented approach when estimating prevalence.

### Results.

### Table 1. Incidences and prevalences worldwide and by region.

| | Natality | Estimated Births | New cases per year | CI lower | CI upper | Last 5 years | Estimated Prevalence |
|---|---|---|---|---|---|---|---|
| **World** | 17 | 137,051,892 | 313,595 | 282,621 | 331,572 | 1,567,975 | 14,343,840 |
| **By region** | | | | | | | |
| **North America** | 11 | 4,125,851 | 9,441 | 8,508 | 9,982 | 47,203 | 431,811 |
| **Central Europe and the Baltics** | 9 | 901,583 | 2,063 | 1,859 | 2,181 | 10,315 | 94,360 |
| **Latin America & Caribbean** | 15 | 9,864,174 | 22,571 | 20,341 | 23,865 | 112,853 | 1,032,384 |
| **Middle East & North Africa** | 20 | 10,166,227 | 23,262 | 20,964 | 24,595 | 116,309 | 1,063,996 |
| **Sub-Saharan Africa** | 34 | 42,836,680 | 98,017 | 88,336 | 103,635 | 490,083 | 4,483,283 |
| **East Asia & Pacific** | 10 | 23,844,636 | 54,560 | 49,171 | 57,688 | 272,800 | 2,495,578 |
| **South Asia** | 18 | 35,127,717 | 80,377 | 72,439 | 84,985 | 401,887 | 3,676,464 |
| **United States** | 11 | 3,818,030 | 8,736 | 7,873 | 9,237 | 43,681 | 399,595 |

### Table 2. Incidences and prevalences by US ethnicity.

| | Natality | Estimated Births | New cases per year | CI lower | CI upper | Last 5 years | Estimated Prevalence |
|---|---|---|---|---|---|---|---|
| **United States** | 11 | 3,818,030 | 8,736 | 7,873 | 9,237 | 43,681 | 399,595 |
| **By ethnicity (% of US population)** | | | | | | | |
| **US White (57.8%)** | 11 | 2,118,006 | 4,846 | 4,368 | 5,124 | 24,232 | 221,670 |
| **US Hispanic or Latina (18.7%)** | 14 | 852,952 | 1,952 | 1,759 | 2,064 | 9,758 | 89,270 |
| **US Black or African American (12.1%)** | 15 | 342,797 | 784 | 707 | 829 | 3,922 | 35,877 |
| **US Asian or Pacific Islander (5.9%)** | 13 | 257,835 | 590 | 532 | 624 | 2,950 | 26,985 |
| **US American Indian or Alaska Native (0.7%)** | 9 | 20,454 | 47 | 42 | 49 | 234 | 2,141 |

**References**


1. Scheffer IE, Zuberi S, Mefford HC, Guerrini R, McTague A. Developmental and epileptic encephalopathies. *Nat Rev Dis Primer*. 2024;10(1):1-19. doi:10.1038/s41572-024-00546-6

2. Oliver KL, Scheffer IE, Bennett MF, Grinton BE, Bahlo M, Berkovic SF. Genes4Epilepsy: An epilepsy gene resource. *Epilepsia*. 2023;64(5):1368-1375. doi:10.1111/epi.17547

3. Karczewski KJ, Francioli LC, Tiao G, et al. The mutational constraint spectrum quantified from variation in 141,456 humans. *Nature*. 2020;581(7809):434-443. doi:10.1038/s41586-020-2308-7

4. Samocha KE, Robinson EB, Sanders SJ, et al. A framework for the interpretation of de novo mutation in human disease. *Nat Genet*. 2014;46(9):944-950. doi:10.1038/ng.3050

5. López-Rivera JA, Pérez-Palma E, Symonds J, et al. A catalogue of new incidence estimates of monogenic neurodevelopmental disorders caused by de novo variants. *Brain*. Published online May 2020. doi:10.1093/brain/awaa051

6. World Bank Open Data. World Bank Open Data. Accessed December 22, 2024. https://data.worldbank.org

7. Martin JA, Hamilton BE, K OMJ, Driscoll AK. Births: Final Data for 2019. Accessed December 22, 2024. https://stacks.cdc.gov/view/cdc/100472

8. Dreier JW, Laursen TM, Tomson T, Plana-Ripoll O, Christensen J. Cause-specific mortality and life years lost in people with epilepsy: a Danish cohort study. *Brain J Neurol*. 2023;146(1):124-134. doi:10.1093/brain/awac042

9. Donnan AM, Schneider AL, Russ-Hall S, Churilov L, Scheffer IE. Rates of Status Epilepticus and Sudden Unexplained Death in Epilepsy in People With Genetic Developmental and Epileptic Encephalopathies. *Neurology*. 2023;100(16):e1712-e1722. doi:10.1212/WNL.0000000000207080

10. CDC. Epilepsy Facts and Stats. Epilepsy. October 3, 2024. Accessed December 23, 2024. https://www.cdc.gov/epilepsy/data-research/facts-stats/index.html

11. Specchio N, Curatolo P. Developmental and epileptic encephalopathies: what we do and do not know. *Brain*. 2021;144(1):32-43. doi:10.1093/brain/awaa371

12. Sanders SJ, Campbell AJ, Cottrell JR, et al. Progress in Understanding and Treating SCN2A-Mediated Disorders. *Trends Neurosci*. 2018;41(7):442-456. doi:10.1016/j.tins.2018.03.011

13. Liu Y, Schubert J, Sonnenberg L, et al. Neuronal mechanisms of mutations in SCN8A causing epilepsy or intellectual disability. *Brain*. 2019;142(2):376-390. doi:10.1093/brain/awy326

14. Chu-Shore CJ, Major P, Camposano S, Muzykewicz D, Thiele EA. The natural history of epilepsy in tuberous sclerosis complex. *Epilepsia*. 2010;51(7):1236-1241. doi:10.1111/j.1528-1167.2009.02474.x
